# Vitamin B12, Homocysteine, and Cognitive Function in the Older Adults Attending Tertiary Geriatric Psychiatry Centre: A Retrospective Case Notes Based Study

**DOI:** 10.1101/2025.08.02.25332580

**Authors:** Ravish Huchegowda, Vijaykumar Harbishettar, Preeti Sinha, Sameeksha Das, PT Sivakumar, John P John

## Abstract

**Background:** Cognitive impairment in the older adult population has been linked to low Vitamin B12 levels and elevated Homocysteine levels, but evidence is not clear. This study examines the relationship between serum Vitamin B12 (B12) levels, Homocysteine (HCY) levels, and cognitive impairment among the Indian older adult population.

**Methods:** This retrospective case notes-based study was conducted on 96 subjects with cognitively normal (n-21), Alzheimers disease (AD; n = 42), and other cognitive disorders (n = 33) with a total sample mean age was 70.1 (+/-8.9) years and 58% male, had cognitive assessment by Hindi Mental Status Examination Scale (HMSE) along with serum B12 levels (pg/ml) and serum HCY levels (micromol/L). Pearsons bivariate correlation, multi-linear regression analysis, and mediation analysis using bootstrapping were done.

**Results:** About 72% had elevated HCY levels (>15 micromol/L) and 33% had low B12 levels (<200 pg/mL). The mean HMSE score was 16.9 (+/-8.5). There was no linear correlation between the HMSE scale and both HCY levels (r = −0.006, p = 0.95) and B12 (r = +0.12, p = 0.23). HCY levels were different between the groups: cognitive impairment (AD: 22.6 +/-13.1 micromol/L; others: 24.0 +/-16.0 micromol/L) and normal group (20.7 +/-13.5 micromol/L), with no statistical significance. The multi-linear regression test results indicated that gender was a significant predictor of the HMSE score, whereas other variables such as B12, HCY, and age did not show significant predictive value contrary to the initial hypothesis.

**Conclusion:** There are no significant independent associations were found between B12 or HCY levels and the cognitive test (HMSE). The data also did not support any statistically significant moderation effect observed between B12 and HCY levels on the HMSE scale.

## Introduction

Vitamin B12 (B12) and Homocysteine (HCY) are important in the cognitive health of the elderly. B12 is important in one-carbon metabolism, its deficiency leads to an increase in HCY levels, leading to neurological damage(1). HCY is recognised as a risk factor for cerebrovascular diseases, dementia(2) and also for Alzheimer’s disease (AD) (3). Elevated HCY causes an increase in oxidative stress, excitotoxicity, and small vessel diseases thereby causing a decline in specific cognitive domains(4).

The epidemiological study showed that significant correlation between high serum HCY levels, low serum B12 levels, and poor cognitive functions(5). Raised HCY is seen as a sensitive marker of B12 and Folate(FLT) deficiency (6). These reports have led to consider that B12 deficiency can lead to cognitive impairment via elevated HCY(7). Following this assumption, several prospective studies have also shown that B12 deficiency and elevated HCY may predict progressive cognitive decline among the ageing population. It is further reported that B12 deficiency may cause reversible cognitive impairment if managed promptly, making it clinically important(8).

However, many studies have not found clear evidence for the above-mentioned associations. While some trials on B12 supplementation in mild deficiency after being corrected have not seen a reversal of cognitive impairment(6). The impact of raised HCY may have more effects on specific cognitive domains, particularly executive function, rather than on global cognition or memory(9). For instance, a recent study reported that a non-demented elderly population of more than 80 years was found to have higher HCY with poorer executive function as compared to global cognition or memory(10). This relationship continued even after correcting B12 and FLT, suggesting that HCY had an independent effect on the frontal executive cognitive aspects(11). These findings suggest a complex relationship confounded by population characteristics, neurological disorders, and cognitive assessment scales used.

B12 deficiency is common in India due to nutritional factors associated with vegetarian diets and malabsorption issues (12,13), while many studies are done in Western countries showing the links between elevated HCY and B12 deficiency causing cognitive decline, such studies in the Indian population remain limited (11). Nutritional deficiencies remain highly prevalent in the Indian population, and some of these can lead to reversible dementia(14). The current study was conducted to explore the association between serum B12 and HCY levels with cognitive function in the Indian older adult population.

## Methods

This was a retrospective electronic medical record (EMR) based analysis. The study was carried out following approval by the XXXXXX Institutional Ethics Committee [Ref No. XXXXXX/IEC (BS & NS DIV) dated 25.01.2024]. Patient records were sourced from the institutional EMR.

### Subjects

All the case notes of the patients visiting the geriatric services in the XXXXXXXX between January 2019 and December 2024, were considered for the study. The Hindi Mental State Examination (HMSE) scores, where available, were noted from the EMR. The data from the biochemical investigation, including serum Vitamin B12 and HCY [by chemiluminescence method (CLIA)], where available, were sourced from the e-hospital database software.

### Sample size estimation

The sample size for this study was estimated based on the anticipated correlation between HMSE scores and serum levels of B12 and HCY. Previous research has suggested a moderate effect size (*r*=0.3) when examining cognitive impairment in relation to biochemical markers (15). To ensure sufficient statistical power (80%) for detecting such associations at a 0.05 significance level, the required sample size was calculated to be approximately 85 subjects.

### Exclusion criteria

EMR records showing major neurological conditions such as Parkinson’s disease, Stroke, and Neuro-infections, notes documented as being on B12 & folate supplementation therapy, and persons exhibiting outlier values for serum B12 (> 2000 pg/ml) were excluded.

### Data analyses and statistics

Case notes from 96 subjects were used for collecting data on cognitive function (HMSE scores), serum B12 levels, serum HCY levels, and demographic details, including age, gender, education, and clinical diagnoses. Biochemical parameters (B12 and HCY levels) were correlated with HMSE scores using Pearson’s correlation coefficient after log transformation of variables for normality. Differences in biochemical parameters between relevant demographic variables, particularly gender, were compared using independent t-tests, and differences across cognitive status groups (normal, Alzheimer’s dementia, and other cognitive disorders) were compared using one-way ANOVA. Multi-linear regression analysis was used to assess the predictive value of B12 and HCY levels on HMSE scores, adjusting for age, gender, and education. Mediation analysis was performed using the Process macro with bootstrapping in IBM SPSS v23 to determine if HCY mediates the relationship between B12 levels and HMSE scores. All statistical analyses were performed using IBM SPSS v23 software, and a p-value less than 0.05 was considered statistically significant.

## Results

### Sample Characteristics

The study sample included 96 participants (56 males, 40 females) with a mean age of 70.1 years (SD = 8.9, range = 52–86). About 68% of participants were aged 60 years or over, with the remainder in their 50 years (the latter were typically persons with early-onset dementias). The average years of education was 8.4 years (SD = 5.2); 25% had no formal education or were illiterate, while 30% had completed high school or above. In terms of residence, 67% hailed from rural areas and 33% from urban areas. Clinically, 96 participants were classified as three groups namely; the cognitively normal group (no diagnosis of impairment; n=21; 21.9%), the Alzheimer’s disease group (AD; n-42; 43.8%), and other cognitive impairments group (n=33; 34.4%)(including 10 with mild cognitive impairment, 15 with vascular dementia, 5 with Frontotemporal dementia/Pick’s disease, and 3 with other psychiatric cognitive disorders).

The study further noted that the normal group had significantly higher HMSE scores than both the AD and other impairment groups, which validated the study sample. The average age of the normal group was slightly younger than that of the AD group, though this difference was not statistically significant (p = 0.09). Gender distribution showed a higher proportion of females in the AD group (50%) compared to the other group (30%), but the differences in gender and education across groups were not significant.

Approximately 40% of the AD group had a B12 deficiency, in contrast to about 24% in the normal group, with no statistical significance (χ^2^ (1) =1.32, p = 0.25). Elevated HCY levels were observed in most participants across all groups, with a non-significant trend suggesting that AD group had lower mean B12 levels and slightly higher HCY levels compared to the normal group. The other impairment group had mean B12 levels like those of the normal group but exhibited the highest mean HCY levels.

Variables: We examined HMSE score as the dependent variable. The following were used as independent variables (predictors): a) log Vitamin B12: Log-transformed B12 level. A natural log transformation was applied to B12 (pg/mL) to reduce skewness. b) log-transformed HCY level. c) log B12 HCY: Log-transformed ratio of B12 to HCY. d) Age in years. e) Years of Education: Formal education duration, in years. f) Gender: Categorical, coded 0 for female and 1 for male.

**Figure 1:**
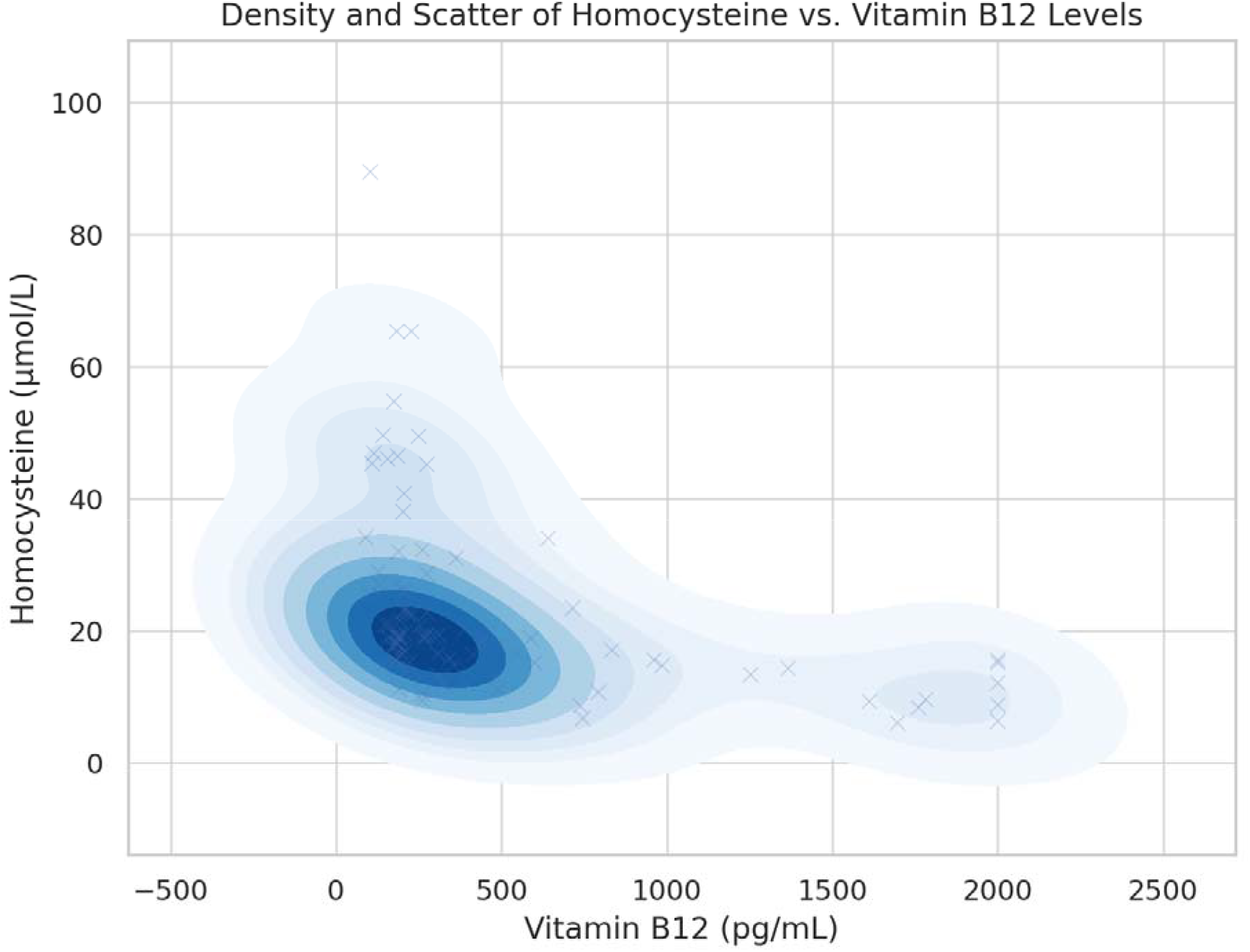
Density and Scatter Distribution of HCY vs. B12 Levels. Figure 1. : Represents a density and scatter plot of HCY (µmol/L) vs. B12 (pg/mL) levels. The contour density shading highlights that most participants cluster around moderate HCY (15-25 μmol/L) and low to moderate B12 levels (200-500 pg/mL), with fewer extreme values observed.

Correlation Analyses: A moderate negative correlation was found between B12 and HCY levels (r = –0.404, p < 0.001), indicating that participants with low B12 tended to have higher HCY levels. Correlation between B12 levels with HMSE scores (r = +0.124), was not statistically significant (p = 0.227). HCY levels had virtually no linear correlation with HMSE scores (r = –0.009, p = 0.95), suggesting that HCY levels do not provide meaningful information about cognitive test scores across the sample.

We also examined correlations within subgroups: Among cognitively normal individuals, B12 and HMSE, neither was significant. These subgroup analyses indicate no clear association between B12/HCY levels and cognitive scores, even when stratified by diagnosis.

Regression Analysis: A multiple linear regression model was run, HMSE was regressed on variables log B12, log HCY, log B12/HCY (ratio), age, education, gender, and the two interaction terms. Variance inflation factors (VIF) and the design matrix condition number were checked to assess multi-collinearity. We also tested for any interaction between B12 and HCY. We included interaction terms to test moderation by gender on the effects of B12 and HCY: specifically, Gender × log B12 and Gender × log HCY were added to the model. The results are summarized in **Table 2**. The overall model was statistically significant [F (5,90) = 4.02, p =.0025)] and accounted for approximately 18.2% of the variance in HMSE scores (R^2^ = 0.182, adjusted R^2^ = 0.137). However, most of this variance was due to gender differences: gender was a significant predictor (β = +0.44, p <.001). Specifically, the coefficient for gender (coded 0 = female, 1 = male) was B = +7.435 (SE = 1.76), indicating that, on average, male participants scored about 7.4 points higher on HMSE than female participants, when controlling for age, B12, and HCY levels. This suggests that in our sample, females had lower cognitive scores than males of similar age and B12/HCY status. This gender effect may reflect the higher prevalence of dementia among women in the sample (as more AD patients were female) rather than a direct biological gender effect.

**Table 1.**
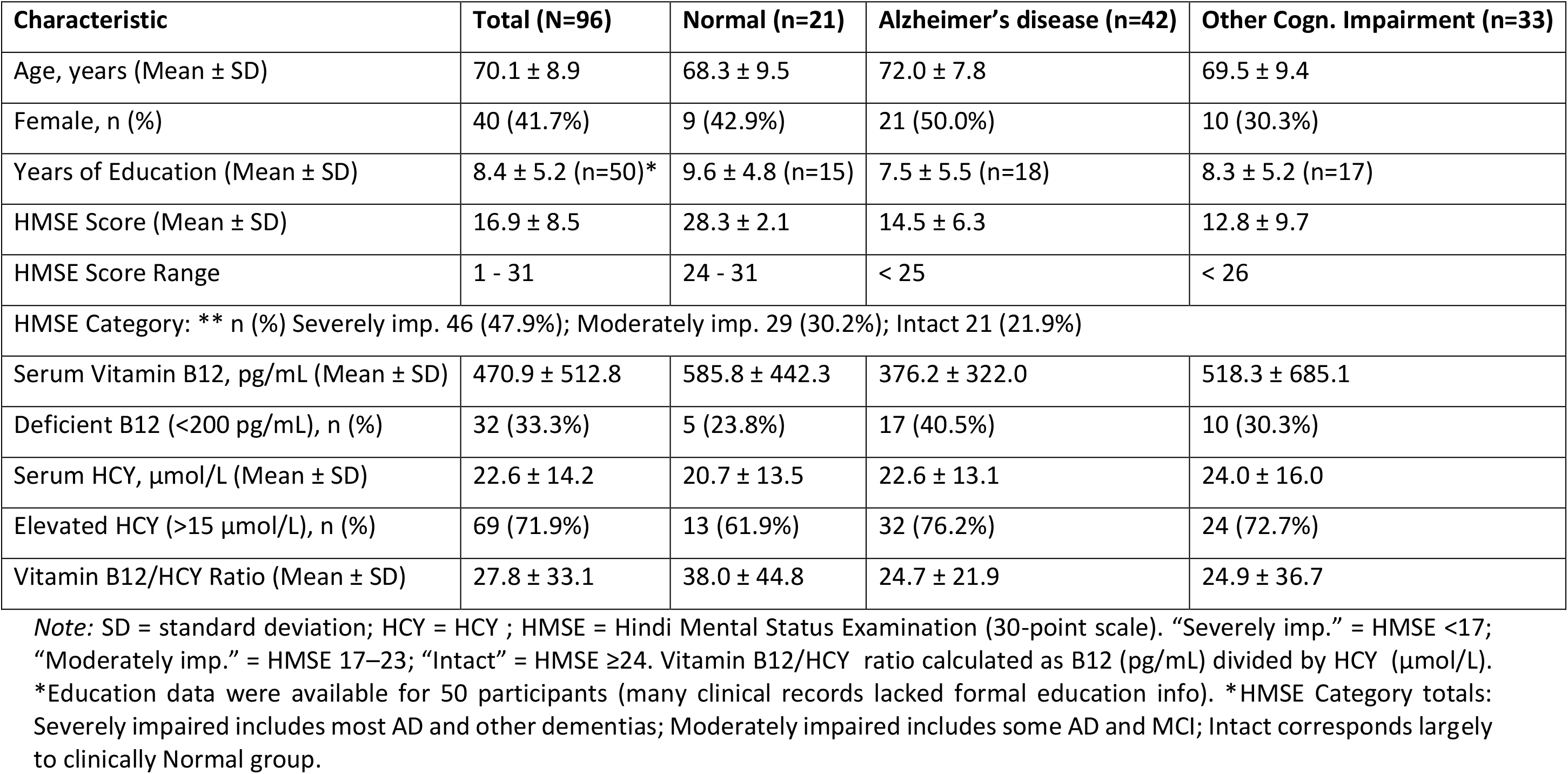
Descriptive statistics for key variables overall and by cognitive status group.

**Table 2.**
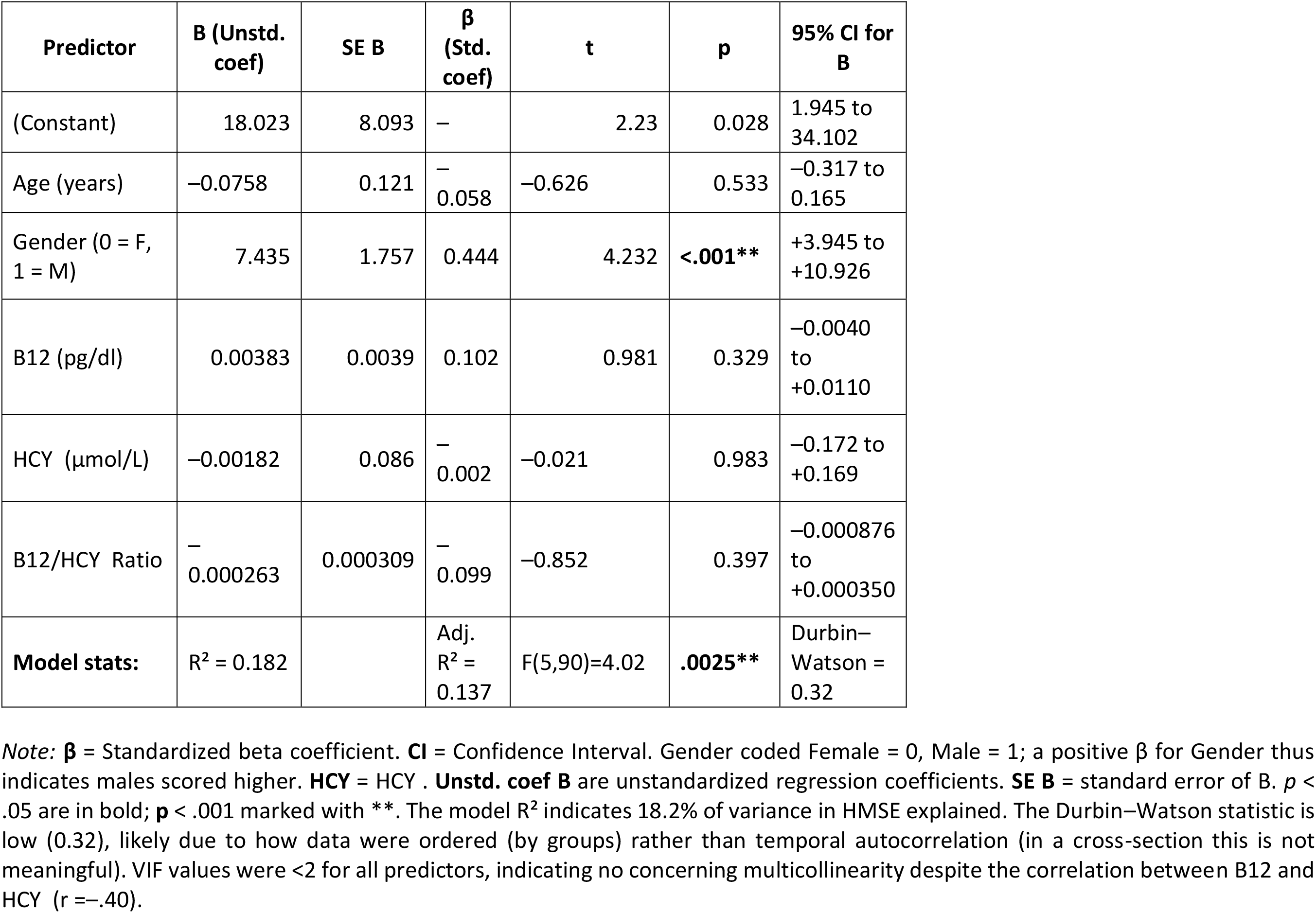
Multi-Linear regression (Stepwise) predicting cognitive score (HMSE) from B12, HCY, and their interaction (with age and gender as covariates).

Importantly, serum B12 level was not a significant independent predictor of HMSE in the regression (B = 0.0038, SE = 0.0039, β = +0.10, p = 0.329). The positive coefficient suggests a trend where higher B12 levels correspond to higher cognitive scores, consistent with the bivariate correlation. However, this effect was small and not statistically reliable after accounting for other factors. Similarly, HCY was not a significant predictor (B = –0.0018, SE = 0.086, β = –0.002, p = 0.983) in the multivariate model. The near-zero coefficient for HCY indicates no linear association with HMSE when controlling for age, gender, and B12. Age also remained non-significant (B = –0.0758, SE = 0.121, β = –0.06, p = 0.533). Notably, in this clinic-based sample, age was not strongly related to HMSE score, likely due to the inclusion of some younger dementia patients and some very old but intact individuals, which reduced a simple age-cognition correlation.

No Interaction (Moderation) Effect: We included an interaction term (Vitamin B12 × HCY) to test for moderation, which determines whether the effect of one biomarker on cognition depends on the level of another biomarker. The interaction term coefficient was negative (B = –0.00026, SE = 0.00031), but it was not statistically significant (p = 0.397). This result indicates that there was no significant interaction between Vitamin B12 and Homocysteine in affecting cognitive scores. The lack of a significant interaction means that our data do not support a strong moderation effect. High Homocysteine did not substantially exacerbate any relationship between low Vitamin B12 and poor cognition beyond their individual (non-significant) effects.

Mediation Analysis: This study investigated whether Homocysteine mediates the relationship between Vitamin B12 and cognitive function (HMSE scores) in elderly individuals using bootstrapping in SPSS via the PROCESS macro by Andrew Hayes. The mediation analysis results indicated that neither Vitamin B12 nor Homocysteine significantly predicted HMSE scores. The direct effect of Vitamin B12 on HMSE was non-significant (B = 0.0024, p = 0.1982), and the indirect effect of Vitamin B12 via Homocysteine was also non-significant (BootLLCI = −0.0025, BootULCI = 0.0013). The overall regression model was weak (R^2^ = 0.0178, p = 0.4343), indicating that Vitamin B12 and Homocysteine explain only a small portion of the variance in cognitive function. Given that the bootstrap confidence intervals for all effects included zero, the results suggest that Homocysteine does not act as a mediator in the relationship between Vitamin B12 and cognition in this sample.

Summary of Mediation Analysis Findings

- No significant mediation effect: Homocysteine does not mediate the association between Vitamin B12 and HMSE scores.
- No direct effect: Neither Vitamin B12 nor Homocysteine serves as a strong predictor of cognitive function.
- Alternative cognitive determinants should be explored: Factors such as education, lifestyle, and comorbidities may have a greater influence on cognitive aging and should be prioritized in future studies.

**Figure 2:**
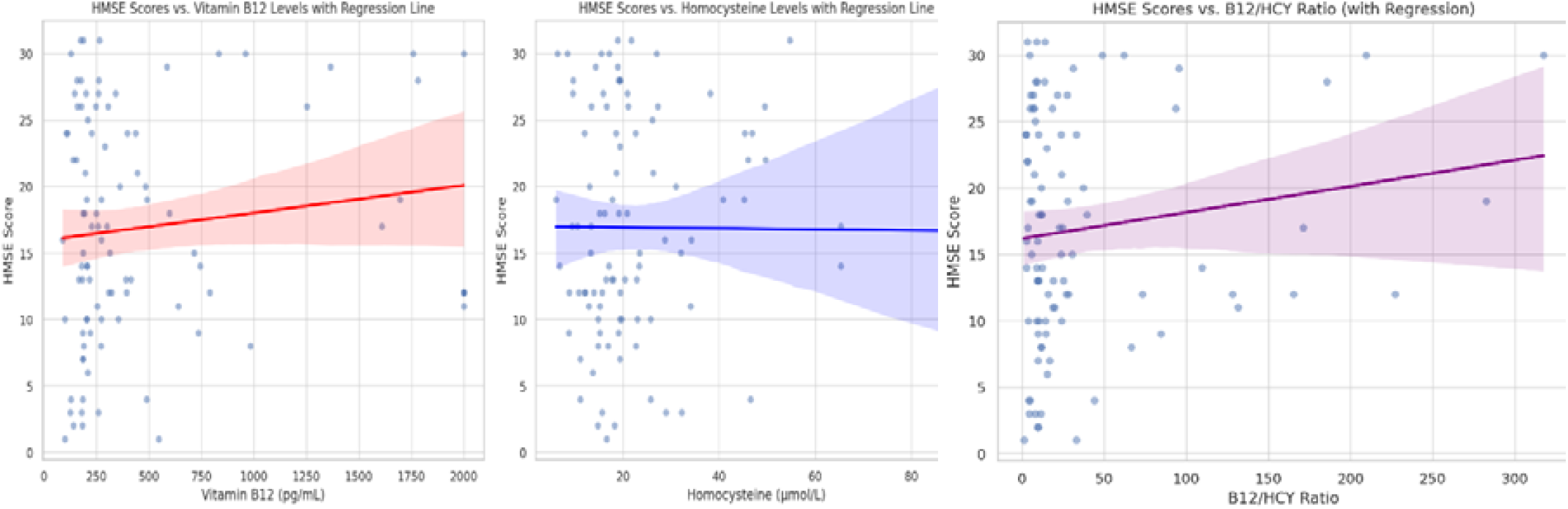
Correlation of HMSE Scores with B12, HCY & B12/HCY Ratio. This figure presents a series of regression analyses examining the correlation between Hindi Mental Status Examination (HMSE) scores and B12, HCY & B12/HCY ratio. The plots show a slight positive trend, indicating that as the B12/HCY ratio increases, HMSE scores tend to improve. However, the wide confidence intervals suggest variability in the association. These findings may imply a potential but weak relationship between cognitive function (HMSE scores) and B12, HCY & B12/HCY biochemical ratio.

## Discussion

Contrary to the authors’ hypothesis, this study found no significant relationship between Vitamin B12, HCY, and the HMSE scale after adjusting for age and gender. Neither Vitamin B12 deficiency nor elevated HCY was found to be an independent risk factor for lower cognitive scores. Additionally, the analysis did not seem to support the concept that HCY moderates the effect of Vitamin B12 on cognition.

The findings from this study must be interpreted in the context of the sample characteristics and existing literature. The sample studied included a mix of cognitively normal individuals and patients with established cognitive impairments such as dementia. Generally, the determinants of cognitive performance are multifactorial (16). Before dementia clinically manifests, a Vitamin B12 deficiency might not significantly lower the cognitive score beyond the impact of the neurodegenerative disease process (17). This could explain why small differences were observed in mean Vitamin B12 levels between dementia patients and normal subjects despite Vitamin B12 levels being lower among AD patients. Notably, the results align with a few previous studies that have not found any relationship between Vitamin B12 levels and global cognition in patients (18, 19). On similar ground, Eussen et al (20) observed that there was no cognitive improvement with Vitamin B12 or FLT in those with mild deficiency. These results are in line with the findings from this study that in a sample with dementia subjects, Vitamin B12 deficiency status by itself did not seem to be a key contributor to cognitive ability.

In contrast, population-based studies of healthy older adults had often found stronger links., such as the Framingham Heart Study (21) (Seshadri et al., 2002) and others showed elevated HCY was associated with an increased risk of developing dementia and cognitive decline over the years (3) (Smith et al., 2018). A meta-analysis by Smith and Refsum (2021) (6) also reported that individuals with AD tend to have higher HCY and lower Vitamin B12 levels than controls. It is possible that low Vitamin B12 and high HCY contribute to the initial development of cognitive impairment as risk factors, but once impairment is present, their association with cognitive test scores can be weak, as observed in this study (22) (Smith et al., 2010).

HCY has been widely implicated in cognitive aging, as it is neurotoxic at higher levels and can induce cerebrovascular damage (3) (Smith et al., 2018). One might expect hyper-homocysteinemia to correlate with worse cognition, and indeed, some studies on non-demented elderly found that higher HCY predicted poorer performance, especially in executive functions (23) (West et al., 2011). This was not the case in this study. One of the reasons could be the cognitive measurement: we used HMSE, a general screening tool mostly that heavily weighs memory, orientation (24) (Bajpai et al., 2020). HCY’s impact might selectively affect executive function and processing speed, which HMSE may not sensitively capture (25) (Zhou et al., 2022). In West et al.’s study (23) (West et al., 2011), HCY correlated with executive-language composite scores but not with memory.

Another reason could be sample characteristics: the very elderly non-demented individuals in prior studies are a different population from ours, which included many dementias. In those without dementia, subtle vascular cognitive impacts of HCY can be detected. But in persons with dementia, cognitive scores are already low and constrained which can be influenced by what is called a “floor effect” obscuring correlations (26) (Yeo et al., 2016). Without longitudinal tracking, it is difficult to conclude whether HCY had a role in causing their condition earlier in life.

The mediation analysis was grounded in the theoretical model that B12 influences cognition via HCY. One interpretation is that B12 may need to be severely deficient before cognitive effects become pronounced (B12-Associated Neurological Diseases, 2025). Most of the “deficient” patients had moderately low B12 (100–200 pg/mL) but not extreme lows, and they may not exhibit frank dementia purely from B12 lack. Those with very low B12 in the sample did have severe cognitive impairment, but they were few in number and often already diagnosed with neurodegenerative dementia, making it hard to attribute causation. It’s also possible that Folate or Vitamin B6 levels (not measured here) could modify the impact of HCY e.g., concurrent Folate deficiency could exacerbate HCY ‘s effects (28). Without data on Folate, the analysis could adjust for that.

One notable finding was the gender effect; women performed worse on the HMSE than men, on average, in our adjusted model. This could be a sampling artifact: perhaps more women in this study had AD which is plausible since women often have higher AD prevalence. Indeed, 50% of the AD group were female vs. only 30% of the “Other” cognitive disorders group. In fact, large population studies generally find no difference in overall dementia prevalence by sex after accounting for age (29) (Huque et al., 2023); some find women have an advantage in verbal memory but worse in visuospatial tasks. Despite controlling for age, gender effect still persisted, reminding us that social factors or health factors could differ by gender (30) (Sundarakumar et al., 2021).

### Clinical Implications

First, this study reinforces the need for Vitamin B12 screening in the elderly, not because it will necessarily explain cognitive status, but because Vitamin B12 deficiency is common and is one of the few reversible causes of cognitive impairment (31)(Mouchaileh, 2023). It is possible that those who were identified as Vitamin B12-deficient were treated and hence avoided worse cognitive outcomes. Second, the high prevalence of Hyper-homocysteinemia (72%) in our sample is noteworthy. This aligns with prior findings that Indian populations, even “apparently normal” elderly, have a high rate of elevated HCY (32) (Kamdi and Palkar, 2013), possibly due to dietary Folate/Vitamin B12 insufficiencies and genetic factors (9) (Sandhya et al., 2024). Addressing high HCY through B12 and folate supplementation along with lifestyle changes, may have benefits in reducing vascular events, which in turn could protect the brain (33) (Pinzon et al., 2023).

### Limitations

The sample was drawn from a single tertiary center with participants from different regions with different lifestyles, which may limit generalizability. The cross-sectional design precludes any causal conclusions or detection of longitudinal effects; a one-time cognitive score may not reflect subtle influences of Vitamin B12/HCY that manifest over the years. The cognitive assessment relied on HMSE, not a comprehensive tool, and might not capture domain-specific impairments; a neuropsychological battery could provide more insight into which cognitive domains (memory vs executive vs attention) might be affected by Homocysteine. The study may appear as incomplete as data on some confounding factors like folate levels, renal function that affect HCY, and Apolipoprotein E (APOE) genotype any of which could influence cognition and interact with B12/HCY was sought (42).

## Conclusion

In conclusion, this research study on Vitamin B12, Homocysteine, and cognitive function in an XXXXX elderly cohort found no significant direct impact of these biochemical factors on global cognitive performance in a mixed group of cognitively normal and impaired individuals. Homocysteine did not emerge as the mediating pathway between Vitamin B12 and cognition in this study, nor did it act as an effect modifier. These findings temper the expectation that correcting mild Vitamin B12 deficiencies or lowering HCY will substantially improve cognitive scores in patients with established cognitive impairment. However, given the high prevalence of Vitamin B12 deficiency and Hyper-homocysteinemia in older adults, especially in XXXXX, the results should not discourage routine nutritional assessments. Instead, these results highlight that prevention and early intervention might be key: maintaining adequate Vitamin B12levels through diet or supplements in midlife could possibly mitigate one risk factor for later cognitive decline, even if intervention late in the disease course shows limited benefit. Understanding the interplay of nutrition with brain aging will continue to be important as we seek multifaceted strategies to preserve cognitive function in our growing elderly population.

## Data Availability

All data produced in this study are available from the corresponding author upon reasonable request.

